# Therapeutic journey of patients affected by stroke: cohort study

**DOI:** 10.1101/2023.10.09.23296782

**Authors:** Marcirene Santos de Mendonça, Caio Lopes Pinheiro de Paula, Douglas dos Santos, Marcelo Nunes da Silva, Maria Júlia Oliveira Ramos, Fernanda Gomes de Magalhães Soares Pinheiro

## Abstract

The therapeutic journey of patients affected by a stroke cause health impacts and deaths. Stroke is an acute neurological dysfunction, classified as hemorrhagic or ischemic. This study aimed to compare the therapeutic journey in the access of patients affected by stroke in hospitals in the Brazilian Northeast.

Prospective cohort study, carried in all public hospitals that had a neuroimaging service in the State of Sergipe. The collection instrument sought sociodemographic characterization and timeline. The data were exported to the R Core Team 2022 software. After being coded and tabulated, they were analyzed using descriptive statistics. The hypothesis of independence was tested using Pearson Chi-Square and Fisher Exact tests.

A total of 159 patients cared for in the hinterland region of the State and 91 in the capital city (Aracaju) participated in the study, with a median age of 66 years old (SD:55.5-75) in the capital and with a median age of 72 years old (SD:60-82) in the hinterland. There was a predominance in females, 76.6% and 64.1%, respectively. It was found a higher incidence of the ischemic stroke (IS) (p<0.002). The decision time between the onset of signs and symptoms and the decision to call up transportation, it was observed that patients with IS cared for in the hinterland take three times longer (p=0.002). Regarding the time between the stroke detection and the CT scan, it was five times longer for those from the hinterland (p<0.001) in cases of IS. In cases of HS, the time was 1.4 longer for those from the hinterland. Concerning the neurological evaluation, in cases of IS and HS, the rates found were 76.6% *vs* 78.3% in the hinterland, while 100% of patients were evaluated for both types of stroke episodes in the capital city (p<0.001).

The therapeutic journey of patients cared for in the hinterland of the State has a longer period of time.

## Introduction

The therapeutic journey of patients affected by a stroke runs through several challenges, especially in terms of accessing the service and specialized network. Stroke is responsible for the second largest cause of clinical deaths in the world. The World Health Organization (WHO) estimates that it will continue at the same level by 2030.^1^ In the United States, the annual incidence of new or recurrent stroke is about 795,000 cases, of which about 610,000 are first-time strokes and 185,000 are recurrent strokes.^1^ In Brazil, the incidence is around 232-344,000 new cases/year, and it is the largest cause of disability in the population aged over 50 years. ^3,4^

In Brazil, the Stroke Care Line Manual recommends that the health system must provide neurological care coverage, available within 30 minutes of the admission of patients (on-site duty, remote on-call, or even specialized neurological support through telemedicine/telehealth). In the treatment of acute stroke, the possibility of intravenous thrombolysis must be evaluated within a window of up to 4h30min from the onset of symptoms. ^5^

The prognosis of patients affected by stroke depends on the speed of care, which implies early treatment after detection of signs and symptoms, which must be understood as an emergency that requires rapid contact with emergency medical services; however, the greater the time elapsed, the worse the neurological predictors. The time between the onset of accident symptoms and diagnosis/treatment is important for reducing mortality and morbidity in populations. Reducing this time is a priority objective in all stroke programs. ^6^

Stroke is a brain disorder that is attributed to a dramatic rupture or blockage of blood vessels within the brain, thereby preventing blood flow to the brain. Stroke can be classified into ischemic and hemorrhagic stroke (HS), both of which cause local hypoxia that damages brain tissue and produce neurological deficits that can be permanent or temporary. ^8^

It is perceived that the timeline and management of the stroke influence the therapeutic journey, that is, the movement of the patients and caregivers in search of care, from the emergence of a problem or disease to its cure, stabilization or death. A study showed that the therapeutic journey of people affected by stroke in the context of the Brazilian Unified Health System (SUS, as per its Portuguese acronym), exposes them to unpleasant experiences, difficulties in terms of accessing services, inaccuracy in terms of recognizing the warning signs of stroke, as well as stressful experiences, such as distress and expectations when waiting for care. ^9^

Data available in the SUS system point to a disparity in information, which leads to underreporting of cases and, consequently, impacts on the resources available to assist these patients. Furthermore, there is a scarcity of established and/or implemented care protocols, where specialized evaluation by a neurology or neurosurgery team works, in some public hospitals, with an opinion or warning scheme, that is, the hospital does not always have this evaluation available for 24 hours, which certainly causes impacts on stroke response time.

The main purpose of the present study is to compare the therapeutic journey in the access of patients affected by a stroke in hospitals in a State in the Brazilian Northeast.

## Methodology

### Study Design and Participants

All patients present in this prospective cohort were affected by stroke. The collection site consisted of all public hospitals in the State of Sergipe that have a neuroimaging service available 24 hours a day, this being a total of three hospitals, one located in the city of Aracaju, Capital City of the State, and the other two in the hinterland of the State, more precisely in the cities of Itabaiana and Lagarto. The data collection took place in the emergency room and critical axis. The collection period was from August 2022 to January 2023. The eligibility criteria were patients aged 18 years or over, males and females, whose onset of stroke signs and symptoms occurred within the territory of Sergipe and with diagnostic confirmation of stroke through tomography and medical evaluation.

Patients who died before the data collection, those who initially received care in a private network, those who had a change in stroke diagnosis during hospitalization and those who were unable to be contacted by the researcher by telephone after five attempts, 30 days after the stroke episode, were excluded. The sampling technique used was non-probabilistic for accessibility and convenience. Once all these criteria were met, and acceptance to participate in the research was noted, the Free and Informed Consent Form was applied for signature.

The collection period began on the first day of August 2022. This process took place through the application of the collection instrument, which addressed the variables, in order to respond to the research objectives, such as socioeconomic and demographic profile, timeline, that is, from the onset of signs and symptoms, the decision to call up transportation, first care, imaging tests, such as computed tomography, stroke detection, as well as evaluation by a neurologist.

This study complies with the STROBE (Strengthening the Reporting of Observational Studies in Epidemiology) guidelines. ^10^ The data was collected daily and stored in a database, where initially four capable and trained researchers completed the filling. In a second moment, it was reviewed by them; and, finally, it was validated by the responsible researcher, with a view to minimizing errors and improving accuracy of the collected data. The conclusions of this study are available from the corresponding author upon reasonable request.

The study was approved on June 7, 2022, by the Research Ethics Committee of Faculdade Estácio de Sergipe - Estácio Phase, with a favorable opinion under number 2.830.187.

### Statistical Analysis

The statistical analysis in this study was carried out using the R Core Team 2022 software (Version 4.2.3), with a significance level of 5%. Descriptive statistics was used. Categorical variables were described using absolute and relative percentage frequencies. Continuous variables were described using mean, median, standard deviation and interquartile range. ^11^

The hypothesis of independence between categorical variables was tested using Pearson Chi-Square and Fisher Exact tests. ^11^ The hypothesis of intra-class adherence of continuous variables and normal distribution was tested using the Shapiro-Wilk test. ^12^ As this was rejected, the hypothesis of equality of medians was tested using the Mann-Whitney test (two groups). ^13^

## Results

### Sociodemographic profile of patients affected by stroke in the State of Sergipe

The sample studied consisted of 250 patients, 91 of whom were treated in Aracaju, Capital City of the State of Sergipe, and 159 were treated in the hinterland of Sergipe. With respect to the clinical profile of these patients, the findings of this study demonstrated a relatively even distribution in the allocation by gender, with a small incidence of females. Concerning age, people affected by this condition generally present themselves as elderly. According to this study, a median age of 66 years old was obtained in the capital city, SD [55.5-75], while, in the hinterland, this median was 72 years old, SD [60-82].

With respect to self-declared race/color, there was a higher incidence of the black race in relation to the white race in both places, and, to a lesser extent, the indigenous race. The variable related to the profession or occupation of patients was relevant (p=0.002), highlighting that, when comparing capital and hinterland, 58.2% *vs* 74.2% were retired, respectively (Table 1).

**Table 1.** Sociodemographic characteristics of patients affected by a stroke in hospitals in the capital (Aracaju) and in the hinterland of the State of Sergipe, 2023.

|  | Origin |  | p-value |
| --- | --- | --- | --- |
|  | Capital | Hinterland |  |
| **Age, Median [IQR]** | 66 [55.5-75] | 72 [60-82] | 0.005<sup>M</sup> |
| **Gender, n (%)** |  |  |  |
| Male | 41 (45.1) | 83 (52.2) | 0.277<sup>F</sup> |
| Female | 50 (54.9) | 76 (47.8) |  |
| **Race, n (%)** |  |  |  |
| White/Yellow | 24 (26.4) | 55 (34.6) | 0.057<sup>CH</sup> |
| Black/Brown | 67 (76.6) | 102 (64.1) |  |
| Indigenous | 0 (0) | 2 (1.3) |  |
| **Social Class, n (%)** |  |  |  |
| 2 to 4 MW | 10 (11) | 17 (10.7) | 0,938<sup>CH</sup> |
| <2 MW | 62 (68.1) | 112 (70.4) |  |
| Non-identified | 19 (20.9) | 30 (18.9) |  |
| **Marital Status, n (%)** |  |  |  |
| Married | 45 (49.5) | 68 (42.8) | 0,594<sup>CH</sup> |
| Stable Union | 7 (7.7) | 15 (9.4) |  |
| Divorced | 9 (9.9) | 11 (6.9) |  |
| Single | 8 (8.8) | 22 (13.8) |  |
| Widowed | 22 (24.2) | 43 (27) |  |
| **Profession or Occupation, n (%)** |  |  |  |
| Self-employed worker | 24 (26.4) | 14 (8.8) | 0.002<sup>CH</sup> |
| Public servant | 2 (2.2) | 2 (1.3) |  |
| Private company employee | 2 (2.2) | 2 (1.3) |  |
| Retired | 53 (58.2) | 118 (74.2) |  |
| Other | 10 (11) | 23 (14.5) |  |
| **Education, n (%)** |  |  |  |
| Elementary school | 55 (60.4) | 77 (48.4) | 0.138<sup>CH</sup> |
| High school | 9 (9.9) | 10 (6.3) |  |
| Higher education | 2 (2.2) | 4 (2.5) |  |
| Post-Graduation | 0 (0) | 1 (0.6) |  |
| Never studied | 25 (27.5) | 67 (42.1) |  |

Caption: IQR – Interquartile Range. n – absolute frequency. % – percentage relative frequency. M – Mann-Whitney test. CH – Pearson Chi-Square test. F – Fisher Exact test.
Source: Prepared by the author (2023).

Regarding income, there was proportionality, both in the capital city (68.1%) and in the hinterland (70.4%), they receive <2 minimum wages (≈ R$2,604.00). Married marital status was more prevalent in the capital city (49.5%), compared to the hinterland (42.8%). As for education, most (60.4%) patients have elementary school in the capital city, while 27.5% have never studied. When compared to the hinterland, it is observed that 48.4% of patients have elementary school *vs* 42.1% who never studied (p <0.138) (Table 1).

### Stroke incidence in the State of Sergipe

As for the classification of stroke as ischemic (IS) and hemorrhagic (HS), both in the capital and in the hinterland, there was a higher incidence of IS, with 76.9% in the capital, compared to 91.2% in the hinterland. For HS, the incidence was 23.1% in the capital vs 8.8% in the hinterland (p <0.002).

### Therapeutic journey of patients affected by stroke in the capital and in the hinterland

Regarding the therapeutic journey of patients treated in the capital and in the hinterland, the variable “time” was compared, being recorded from the onset of the stroke signs and symptoms until the diagnostic conclusion, as well as the neurological evaluation. When describing this variable, it was possible to associate it from the movement of patients from the place where the stroke signs and symptoms began to the place of diagnostic confirmation (Figure 1).

**Figure 1.**
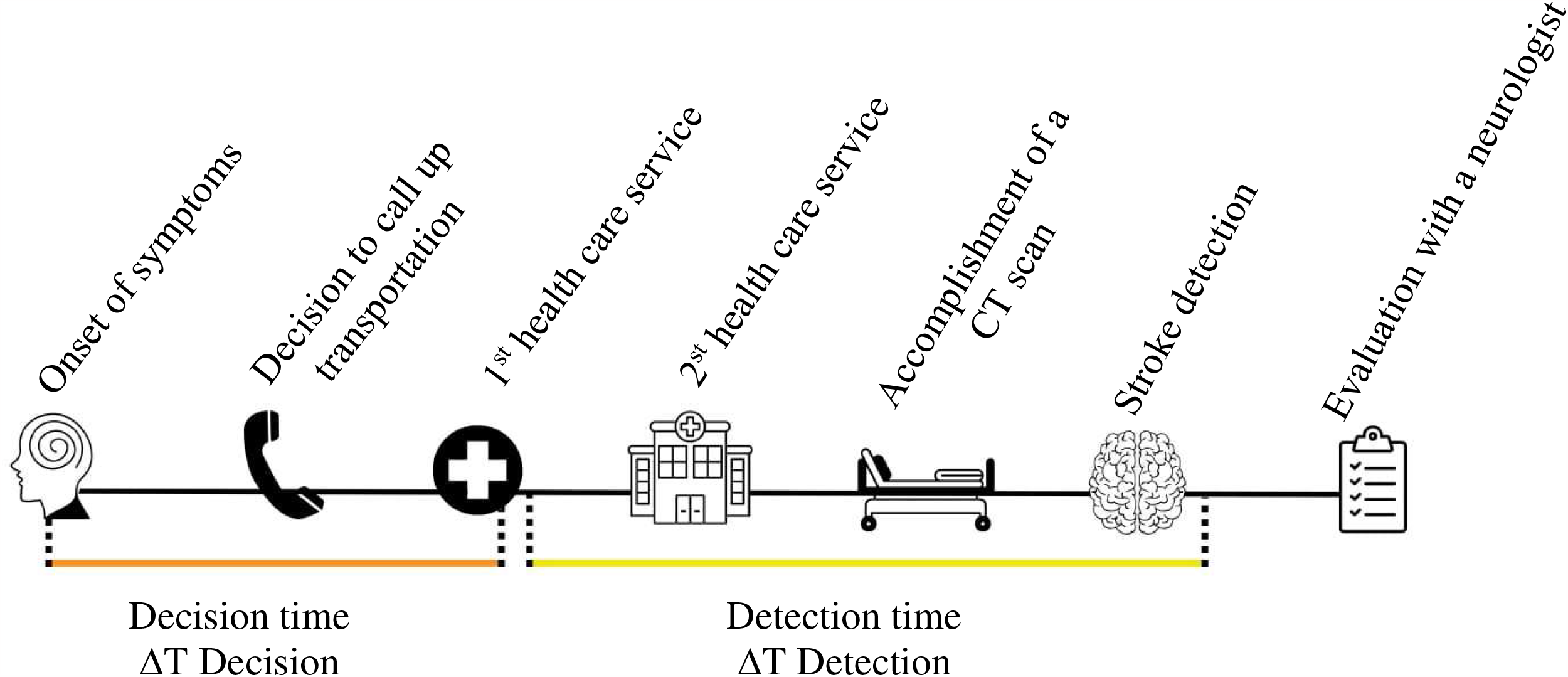
Timeline from the onset of signs and symptoms to the stroke diagnosis and subsequent neurological evaluation. Source: Prepared by the author (2023).

When checking the means of transportation used by patients until their arrival at the first care, a significant difference was obtained (p = 0.053). Patients with stroke treated in the capital city traveled using their own transportation (48.6%), Mobile Emergency Care Service (SAMU, as per its Portuguese acronym) (34.3%) and other means of transportation (17.1%), such as an ambulance provided by the City Hall, transportation app and others. Patients with stroke treated in the hinterland traveled using their own transportation (57.9%), SAMU (19.6%) and other means of transportation (22.5%).

## DECISION TIME (∆T DECISION) IN IS

As for the analyzed times, in the capital and in the hinterland, patients with IS had the following results (Table 2):

**Table 2.** Time elapsed and factors that influence the search for medical care by those affected by a stroke in hospitals in the capital (Aracaju) and in the hinterland of the State of Sergipe, 2023.

| Time and factors | IS |  | p-value | HS |  | P-value |
| --- | --- | --- | --- | --- | --- | --- |
|  | Capital | Hinterland |  | Capital | Hinterland |  |
| Time between the onset of symptoms and the decision to call up transportation (h), *Median [IQR]* | 0.8 [0.2-3] | 2.5 [0.5-13.5] | 0.002<sup>M</sup> | 0.5 [0.2-1] | 5.2 [0.5-72] | 0.028<sup>M</sup> |
| Time between the decision to call up transportation and the first care (h), *Median [IQR]* | 1.5 [0.7-2.5] | 1.5 [0.8-2.9] | 0.673<sup>M</sup> | 0.8 [0.5-2] | 1 [0.4-2] | 0.934<sup>M</sup> |
| Time between the onset of symptoms and their arrival at the care unit that offers a neuroimaging service, *Median [IQR]* | 9.6 [4-27.2] | 8.4 [3.4-25.9] | 0.720<sup>M</sup> | 7.7 [5-15.9] | 12.2 [2.6-80.5] | 0.561<sup>M</sup> |
| Time between the first care and the accomplishment of a CT scan (h), *Median [IQR]* | 1 [0.6-2.3] | 2 [0.7-5.5] | 0.009<sup>M</sup> | 1 [0.5-4.6] | 3 [1.5-5.5] | 0.077<sup>M</sup> |
| Time between the CT scan and the stroke detection (h), *Median [IQR]* | 0.7 [0.3-2.5] | 3.6 [1-19.6] | <0.001<sup>M</sup> | 0.5 [0-1.5] | 0.7 [0-2.7] | 0.829<sup>M</sup> |
| Time between the onset of symptoms and the stroke detection (h), *Median [IQR]* | 18.1 [7-37.5] | 25 [13.5-59.2] | 0.007<sup>M</sup> | 11.3 [7.7-25.5] | 19.9 [9.3-85] | 0.325<sup>M</sup> |
| Time between the onset of symptoms and the stroke detection, *n (%)* |  |  |  |  |  |  |
| <4:30 | 12 (17.1) | 8 (5.5) | 0.011<sup>F</sup> | 4 (19) | 2 (14.3) | 1.000<sup>F</sup> |
| >4:30 | 58 (82.9) | 137 (94.5) |  | 17 (81) | 12 (85.7) |  |
| Neurological evaluation, *n (%)* | 70 (100) | 111 (76.6) | <0.001<sup>F</sup> | 21 (100) | 11 (78.6) | 0.056<sup>F</sup> |

Caption: IQR – Interquartile Range. n – absolute frequency. % – percentage relative frequency. M – Mann-Whitney test. F – Fisher Exact test.
Source: Prepared by the author (2023).

### Time between the onset of symptoms and the decision to call up transportation

The median time in the capital is 0.8 hours [IQR: 0.2-3] *vs* 2.5 hours [IQR: 0.5-13.5] in the hinterland (p=0.002).

### Time between the decision to call up transportation and the first care

The median time is proportional in the capital and in the hinterland, 1.5 hours [IQR: 0.7-2.5] and 1.5 hours [IQR: 0. 8-2.9], respectively (p=0.673).

### Time between the onset of symptoms and their arrival at the care unit that offers a neuroimaging service

The median time in the capital is 9.6 hours [IQR: 4-27.2] *vs* 8.4 hours [IQR: 3.4-25.9] in the hinterland (p=0.720).

## DETECTION TIME (∆T DETECTION) IN IS

### Time between the first care and the accomplishment of a CT scan

The median time in the capital is 1 hour [IQR: 0.6-2.3] *vs* 2 hours [IQR: 0.7-5.5] in the hinterland (p=0.009).

### Time between the CT scan and the stroke detection

The median time in the capital is 0.7 hours [IQR: 0.3-2.5] *vs* 3.6 hours [IQR: 1-19.6] in the hinterland (p=0.001).

### Time between the onset of symptoms and the stroke detection

The median time in the capital is 18.1 hours [IQR: 7-37.5] *vs* 25 hours [IQR: 13.5-59.2] in the hinterland (p=0.007).

### Time between the onset of symptoms and stroke detection (less than 4 hours and 30 minutes)

This time interval in the capital was 17.1% *vs* 5.5% in the hinterland (p=0.011).

### Neurological evaluation

In the capital, all cases (100%) were evaluated by neurologists, while the evaluation rate in the hinterland was 76.6% (p=0.001).

## DECISION TIME (∆T DECISION) IN HS

As for hemorrhagic stroke, one can observe the following findings between the capital and the hinterland (Table 2):

### Time between the onset of symptoms and the decision to call up transportation

The median time in the capital is 0.5 hours [IQR: 0.2-1] *vs* 5.2 hours [IQR: 0.5-72] in the hinterland (p=0.028).

### Time between the decision to call up transportation and the first care

The median time in the capital is 0.8 hours [IQR: 0.5-2] *vs* 1 hour [IQR: 0.4-2] in the hinterland (p=0.934).

### Time between the onset of symptoms and their arrival at the care unit that offers a neuroimaging service

The median time in the capital is 7.7 hours [IQR: 5-15.9] *vs* 12.2 hours [IQR: 2 .6-80.5] in the hinterland (p=0.561).

## DETECTION TIME (∆T DETECTION) IN HS

### Time between the first care and the accomplishment of a CT scan

The median time in the capital is 1 hour [IQR: 0.5-4.6] *vs* 3 hours [IQR: 1.5-5.5] in the hinterland (p=0.077)

### Time between the CT scan and the stroke detection

The median time between capital and hinterland is proportional, 0.5 hours [IQR: 0-1.5] and 0.7 hours [IQR: 0-2.7], respectively (p=0.829).

### Time between the onset of symptoms and the stroke detection

The median time in the capital is 11.3 hours [IQR: 7.7-25.5] and *vs* 19.9 hours [IQR: 9.3-85] in the hinterland (p=0.325).

### Neurological evaluation

In the capital, all cases (100%) were evaluated by a neurologist, while the evaluation rate in the hinterland was 78.6% (p=0.056).

## Discussion

In the present study, females predominated in terms of people affected by stroke, with a median age greater than 60 years old. These data corroborate a study carried out in Lebanon, where the female gender was predominant (53.8%) and the mean age for stroke was 70 years old, regardless of etiological classification. Other Brazilian studies also show it, such as a study carried out in Salvador, which showed a predominance of females (52.7%) and a mean age over 60 years old. ^15,16^ Another study also found a higher incidence of elderly people, with ages ranging between 24 and 100 years old, with a mean of 69 years old. ^17^ These results show that being over 60 years of age is an important risk factor for the occurrence of stroke.

Regarding the classification of stroke, this study shows a higher incidence of IS, both in the capital (76.9%) and in the hinterland (91.2%). As the type of stroke, the findings corroborate national and international studies in which the rate is around 80% in ischemic cases. ^6,18,19,20,21,22^.

With respect to the therapeutic journey of patients affected by stroke, several times were analyzed, dichotomized into decision time (∆T decision) and detection time (∆T detection), as well as evaluation by the specialist neurology team. When comparing the decision time between the onset of signs and symptoms and the decision to call up transportation, it was observed that patients with IS treated in the hinterland spent three times more time compared to those in the capital. This delay is even greater in hemorrhagic cases, where it took ten times longer in the hinterland than in the capital.

In a study in a European country, Spain, patients affected by IS had a median delay of 60 minutes in terms of deciding to call up transportation. ^23^ It is possible to imagine that this delay in terms of seeking help for care is related to the difficulty in relation to recognizing stroke signs and symptoms, in addition to the limited knowledge that it is a medical emergency.

When considering the time between the decision to call up transportation and the first care, the capital and the hinterland had proportional times, with medians of 1.5 hours. In a study carried out in five regions of Brazil, it was observed that travel time is more than two hours, so that the journey from the location of the patient to the place of medical care is a geographical barrier. ^24^

In a cross-sectional study carried out in Europe, the data were divergent, as the interval between the decision to call up transportation and the arrival at the place of medical care had a median of 56 minutes. ^23^ It is possible that care times vary, as there are many geographical barriers in Brazil. In our study, even though it was carried out in the smallest Brazilian State, access to specialized services does not seem to have a geographical distribution that favors better time and resolution, which makes the search for care a saga.

As for the decision time from the onset of signs and symptoms to the arrival of the patient at a place of qualified care, that is, one that has a skull computed tomography as an imaging test. The present study reveals that most patients with stroke, regardless of etiological classification, arrived with a median time of more than 8 hours. These data confirm longer times, such as that of the cross-sectional study by Brandão, ^25^ conducted in the State of Bahia, where this time was around 12 hours. In another documentary analysis, carried out in the State of Ceará, the time between the onset of neurological complaints and the arrival at the reference unit was predominantly longer than 4 hours and 30 minutes. ^17^

The literature indicates, in a retrospective review carried out in the United States, a mean time of 15 hours. ^26^ Another cross-sectional study held in Dakar, Capital City of Senegal, showed a median time of 8.5 hours. ^27^ In a study carried out in Romania, 31.6% of patients arrived at the hospital within 4.5 hours after the onset of the stroke, 4.4% in time intervals between 4.5 and 6 hours after the onset and 28.7% arrived at the hospital more than 24 h after the onset of symptoms. ^28^ On the other hand, a prospective cross-sectional survey carried out in Lebanon showed that only 17.7% arrived late, taking more than 3 hours to the health care unit. ^14^

The discussions regarding time are delicate, because it was evident that investigation designs, materials and methods bring different interpretations for many of the comparisons shown, so that, when investigating time, it is perceived that the cutoff value used to exclude time data pre-hospital care of patients who arrive at the hospital with prolonged delays are different.

With respect to the detection time (∆T Detection), it is worth highlighting that this referred to the time spent in the hospital environment. The interval between the first care and the accomplishment of a CT scan was compared, that is, the time in relation to the imaging test. The patients from the hinterland region take twice as long compared to those from the capital city. Corroborating a Brazilian study, carried out in the South of the country, which obtained a median time of 1.3h (0.76h to 2.26h).^16^ Nonetheless, it was different in international studies, such as one held in Indonesia, where the median was 45 minutes, ^29^ and in another developed in Lebanon, where 32.9% underwent a CT scan in less than 20 minutes. ^14^

The time spent between the CT scan and the detection of ischemic stroke in the hinterland region was five times longer than in the capital city. In cases of HS, the hinterland took 1.4 times longer than the capital. Guidelines show that, for the early treatment of patients with stroke eligible for the use of thrombolytics, the CT scan must be performed within 25 minutes of arrival at the emergency room, and it is also recommended that a neurologist responsible for the care is required to be able to interpret the CT test; however, in its absence, the telemedicine program must be used to assist in the evaluation of neuroimaging in real time. ^30^ It is worth emphasizing that there is a lack of data in the pertinent literature regarding protocols with specific times for patients with HS.

Therefore, it is inferred that the absence of the neurology team or professionals trained to interpret and understand neuroimaging on a 24-hour basis has a direct impact on the diagnostic conclusion. It is worth emphasizing that the hinterland region of Sergipe does not have a neurologist or radiologist available 24 hours a day to provide a diagnostic conclusion. This could possibly have had a direct impact on all patients who were able to undergo thrombolytic treatment. Nonetheless, this was not accomplished.

Another relevant data demonstrated is the time elapsed between the onset of signs and symptoms and the stroke detection. In the capital, the median time is shorter, around 18 hours, while this median is 25 hours in the hinterland. By dichotomizing this time into <4:30 and >4:30 hours, a much higher percentage was obtained, around 80 to 90% of patients arrive with a time >4:30 hours, and a minority with a time <4:30 (capital 17.1% of cases and hinterland 5.5% of cases).

The Stroke Care Line Manual in Brazil proposes that, in cases of IS, the patient must be evaluated for the possibility of intravenous thrombolysis within 4h30min of the onset of symptoms. ^31^ According to the study, this reality is discrepant, due to pre-hospital and in-hospital delays, resulting in conservative treatment.

Regarding the specialized evaluation with a neurologist in the capital, all patients had an evaluation by such a professional. In the hinterland, the evaluation rate was 76.6%. Ordinance Nº 800 in relation to the Stroke Care Line recommends that the hospital must provide neurological care coverage, available within 30 minutes of the admission of patients (on-site duty, remote on-call, or even specialized neurological support through telemedicine/telehealth). ^5^ It is also worth highlighting that the SUS system must offer universal support for everyone; however, it is possible to infer the fragility of this health system, which possibly results in unassisted patients.

As for the movement of patients to carry out their journey, the means of transportation used by them was analyzed. It is observed that the fact of owning a car was the most used resource, followed by SAMU. Similar data were found in two cross-sectional studies. ^15,16^ Own car was the first option of choice due to ease of contact; and, according to verbal reports from patients, the car was used due to difficulties in terms of contacting the emergency service.

In conclusion, this cohort highlights the need for urgent measures to improve not only stroke awareness, but also health care protocols, with the purpose of providing timely and appropriate care to patients with stroke. Given the results found, it is suggested that institutional measures are adopted, such as flows, protocols, organized health care systems and even the implementation of stroke units equipped with CT scanners, access to telemedicine programs and specialized teams, with recognition of the patients and administration of treatments accurately and efficiently.

Furthermore, it is opportune to carry out more studies about this topic, using computational geoprocessing techniques capable of contributing to stroke mapping, risk evaluation, planning and health action evaluation. Such measures can efficiently reduce delays in care and treatment, resulting in better outcomes.

## Data Availability

I, Marcirene Santos de Mendonça, declare that I am available to provide all necessary data to Revista Stroke as requested. I commit to making all relevant information available as quickly as possible, ensuring its quality and accuracy. I am aware of the importance of data for the development of research, and I recognize that my collaboration is fundamental to its success. Therefore, I am at your disposal to provide any necessary information, as well as to answer any questions or additional requests. I further declare that all data provided is reliable and corresponds to reality, being in compliance with the laws and regulations applicable to the protection of personal data and privacy. I assume full responsibility for the security and confidentiality of the data provided, committing myself not to share it with third parties without due authorization or specific purpose previously agreed. I am aware that if there is any change in my data availability, I must immediately inform Revista Stroke.

## Acknowledgements

None

## Funding source

This research did not receive any specific grant from public, commercial or non-profit funding agencies.

## Publications

None

## Notes

### Competing Interest Statement

The authors have declared no competing interest.

### Clinical Trial

None

